# A nanopore-based amplicon-sequencing panel to determine the origin and zoonotic host species of *Plasmodium knowlesi* infections

**DOI:** 10.64898/2026.10.16.26365752

**Authors:** Gustavo da Silva, Parsakorn Tapaopong, Aurel Holzschuh, Jetsumon Sattabongkot, Wang Nguitragool, Cristian Koepfli

## Abstract

Population genomic studies have revealed strong regional structure in *Plasmodium knowlesi* across Southeast Asia. To facilitate routine genomic surveillance, we developed a targeted amplicon sequencing panel that preserves the large- and small-scale population genetic structure. We screened published genomes for short, highly polymorphic targets and a 30-SNP barcode. We validated multiplexed amplification and sequencing using a portable Oxford Nanopore Technologies MinION sequencer on 28 clinical infections from four southern Thai provinces collected between 2018–2023. Twenty-seven loci yielded sequence data and 23 infections with data from at least eight loci were retained for analysis. Across the 23 Thai infections and 104 published Malaysian genomes, 586 diverse SNPs were observed. The amplicon panel and 30-SNP barcode retained the major population structure observed in genome-wide data, including a clear distinction between Bornean and Peninsular Malaysian infections and two clusters in Borneo. Across newly sequenced Thai infections, 25 loci were diverse and showed substantial diversity (mean expected heterozygosity = 0.69). All infections were effectively monoclonal. Pairwise identity-by-descent revealed substantial recent relatedness, with 46 of 253 pairs at or above 0.5. Two Thai genetic groups (n = 12 and n = 11) were observed, mirroring previously observed structure in peninsular Malaysia. Groups were not significantly associated with province or collection year. In conclusion, targeted sequencing can reveal geographically informative population structure across scales and thus support surveillance.

## Introduction

Zoonotic malaria caused by *Plasmodium knowlesi* is an emerging public-health threat in Southeast Asia. The parasite is maintained in macaque reservoirs, particularly long-tailed (*Macaca fascicularis*) and pig-tailed (*Macaca nemestrina*) macaques and transmitted by mosquitoes of the *Anopheles leucosphyrus* group. Infection risk is shaped by forest loss, fragmentation, and forest-edge land use that increase contact among people, macaques, and exophagic vectors (Johnson et al., 2024; Moyes et al., 2016; Shearer et al., 2016; Tobin et al., 2024; van de Straat et al., 2022). *P. knowlesi* has become the dominant cause of malaria in Malaysia and is increasingly recognized elsewhere in the region, including Thailand (Cooper et al., 2020; Hussin et al., 2020; Tapaopong et al., 2024). Human infections can infect *Anopheles dirus* experimentally, but sustained human-mosquito-human transmission has yet to be demonstrated. Current epidemiologic evidence remains most consistent with predominantly zoonotic transmission (Fornace et al., 2023; Kumpitak, 2024).

Clinical and diagnostic features of *P. knowlesi* complicate control and surveillance. *P. knowlesi* has a short, 24-hour asexual cycle and can progress rapidly to high parasitemia and severe disease (Barber et al., 2021; Chang, 2023; Grigg et al., 2018). Available parasite lactate dehydrogenase (pLDH)-based rapid diagnostic tests show substantial variation in sensitivity for *P. knowlesi* (Barber et al., 2013; Grigg et al., 2021; Grigg et al., 2014; Tan et al., 2022). Cross-reactivity with *P. vivax*-pLDH targets can further complicate species identification (Grigg et al., 2014; Tan et al., 2022). Microscopy frequently misclassifies *P. knowlesi* because its blood-stage morphology overlaps with *P. malariae*, *P. falciparum*, and other *Plasmodium* species (Grigg et al., 2021; Huber et al., 2023; Romero et al., 2026). Molecular confirmation is therefore important for surveillance.

Genomic epidemiology allows for insights into transmission chains and population structure. In Malaysian Borneo, whole-genome sequencing (WGS) showed that two sympatric yet divergent *P. knowlesi* populations circulate, which are associated predominantly with long-tailed macaques (*M. fascicularis*-associated; Mf-Pk) and pig-tailed macaques (*M. nemestrina*- associated; Mn-Pk) (Divis et al., 2015; Divis et al., 2018). A third major lineage, the Peninsular Malaysia-associated population (Pen-Pk), predominates in Peninsular Malaysia and is clearly differentiated from the two Bornean populations (Divis et al., 2017; Hocking et al., 2020; Turkiewicz et al., 2023). WGS of 28 Peninsular Malaysian clinical infections suggested additional structure within this region with three subclusters (Pen A, Pen B, and Pen C). Pen C was strongly separated from the other subclusters, and Pen C infections were highly related across large genome segments, whereas Pen A and Pen B were both diverse and were less differentiated, with differentiation concentrated in a limited number of genomic regions (Hocking et al., 2020).

WGS directly from *P. knowlesi* clinical infections remains technically demanding because parasite DNA can represent only a small fraction of total DNA in whole-blood specimens, requiring leukocyte depletion, parasite enrichment, or selective whole-genome amplification which add complexity and cost (Diez Benavente et al., 2019; Oresegun et al., 2022; Pinheiro et al., 2015). Multiplex amplicon sequencing (AmpSeq), which targets multiple predefined genomic regions in parallel, overcomes these limitations and is now established in *P. falciparum* and *P. vivax* (de Cesare et al., 2024; Holzschuh et al., 2023; Kattenberg et al., 2022; Kleinecke et al., 2025; LaVerriere et al., 2022; Siegel et al., 2024). The portability and comparatively low infrastructure requirements of nanopore sequencers can facilitate malaria genomic surveillance in laboratories without access to centralized high-throughput sequencing facilities (de Cesare et al., 2024; Holzschuh et al., 2024; Mwenda et al., 2026). No multiplexed *P. knowlesi* AmpSeq framework has been developed for routine population-genetic surveillance.

Here, we developed and validated a targeted AmpSeq panel for *P. knowlesi* using clinical infections from southern Thailand. We quantified local diversity and relatedness, tested whether variation across the markers retained broad population structure defined by Malaysian WGS genomes and finer peninsular substructure, and evaluated a compact 30-SNP representation of broad regional differentiation.

## Materials and Methods

### Study design, setting, and ethics

This study was approved by the relevant institutional ethics committees (see Ethical approval and consent to participate, below). All laboratory analyses used de-identified DNA archived at the Mahidol Vivax Research Unit, Faculty of Tropical Medicine, Mahidol University, Bangkok, Thailand.

### Panel development

Amplicon target discovery was based on published whole-genome-derived SNP data from 28 Peninsular Malaysian clinical *P. knowlesi* infections described by Hocking et al. (2020; Supplementary Table 2). These infections provided >5,000 SNPs distributed across the 14 nuclear chromosomes for the original marker screen. To identify short regions with dense, common variation suitable for resolving infection diversity and relatedness, we scanned the SNP dataset in 200-bp non-overlapping windows. Within each window, multi-SNP alleles were concatenated into haplotypes and haplotype diversity (Hd) was estimated in R from the observed haplotype frequencies. Windows containing ≥10 SNPs and Hd ≥0.75 were retained, yielding 80 high-diversity candidate regions (mean Hd approximately 0.9).

A subset of the high-diversity windows was taken forward for primer design together with five putative antimalarial resistance-gene orthologues (*pkcrt*, *pkdhfr-ts*, *pkmdr1*, *pkk13*, and *pkdhps*), yielding 43 candidate amplicons in total (Supplementary Tables 3 and 4). Amplicons were designed to remain short (<400 bp; candidate sizes 266-381 bp), with primer melting temperatures in a narrow range for multiplexing. Primer candidates containing long homopolymers or predicted secondary structures were excluded, and primer sequences were screened *in silico* against available *P. falciparum*, *P. vivax*, and human reference genomes to minimize potential cross-reactivity. The chromosomal distribution of all 43 candidates is shown in Supplementary Figure S2.

Forty-three candidate primer sets were tested individually by droplet digital PCR (ddPCR) to verify amplification efficiency, and specificity was evaluated by quantitative PCR (qPCR) using genomic DNA from *P. falciparum* and *P. vivax*. Thirty primer pairs showed robust amplification and were selected for the experimental multiplex panel. These 30 loci represented 13 of the 14 *P. knowlesi* chromosomes; the only chromosome 9 candidate (PKNH_0919000) failed singleplex screening and was not carried forward.

### Thai clinical infections

Twenty-eight clinical infections were represented by archived blood specimens from four provinces of southern Thailand: Songkhla (n = 10), Narathiwat (n = 2), Surat Thani (n = 1), and Yala (n = 15) (Supplementary Figure S1; Supplementary Table 1). Blood specimens were obtained from symptomatic patients aged 15 to 80 years from 2018 to 2023. Species confirmation used nested PCR targeting the 18S rRNA locus following established *P. knowlesi* molecular-diagnostic approaches (Ngernna et al., 2019). Parasite DNA was quantified by ddPCR. Quantification outputs in genome copies per microliter were used to guide input volumes for library preparation and to stratify expected locus recovery. ddPCR used the Bio-Rad EvaGreen Supermix with primers 5′-GGCCATTCGGAATTTAGAG-3′ (forward) and 5′-AGGTCCTTGTTATAGGTGAA-3′ (reverse) targeting PKNH_1218400, a single-copy gene. Cycling conditions were 3 min at 95°C followed by 45 cycles of 95°C for 30 s, 53.5°C for 30 s, and 72°C for 1 min; followed by 72°C for 5 min.

After locus-recovery quality control (≥8 successfully genotyped loci), 23 infections and 27 markers were retained for population-genetic analyses. Infections covered multiple years and provinces: Songkhla (n = 6), Narathiwat (n = 2), Surat Thani (n = 1), and Yala (n = 14).

### ddPCR-AmpSeq library preparation and Oxford Nanopore Technologies sequencing

To support highly multiplexed amplification from low-density infections and to reduce inter- amplicon competition, each reaction was partitioned into approximately 15,000 droplets. After partitioning, most droplets contain zero or one DNA template per locus. Although few droplets will contain template DNA from several loci, competition of multiple loci during PCR amplification is largely reduced (Holzschuh et al., 2023). For each sample, 2 µL of DNA served as input. Reaction mix specifications are given in Supplementary Table 7. Droplets were generated using the Bio-Rad QX200 manual droplet generator. To suppress non-specific amplification in the multiplexed reaction, a touchdown PCR profile was used. The annealing temperature started 20 °C above the final annealing temperature (73 °C) and decreased by 1 degree C per cycle to 53 °C, followed by 35 cycles at 53 °C. After thermocycling, droplets were destabilized to recover pooled amplicons. The emulsion was mixed with 70 µL of chloroform and 30 µL of TE buffer, vortexed for 1 min, and centrifuged at 16,300 × g for 10 min to separate phases. The upper aqueous layer of approximately 30 µL containing the amplicons was transferred and purified with magnetic beads. DNA was eluted in 20 µL of elution buffer.

Barcoded libraries were prepared using the Oxford Nanopore Technologies Native Barcoding Kit 96 V14 (SQK-NBD114.96). End-repair and dA-tailing were performed using the NEBNext Ultra II End Repair/dA-Tailing Module (New England Biolabs), followed by barcode and adapter ligation using NEB T4 DNA ligase according to Oxford Nanopore Technologies ligation guidelines. Magnetic bead cleanup was performed between enzymatic steps. Libraries were quantified using the Qubit dsDNA High Sensitivity Assay (Thermo Fisher Scientific), equimolarly pooled, and loaded onto MinION R10.4.1 flow cells for sequencing.

### Haplotype calling and quality control

Raw POD5 data were basecalled with Dorado v1.1.1 using the super-accuracy model. Adapter trimming and demultiplexing were performed in Dorado. Only reads with Phred quality ≥ Q20 were retained to reduce sequencing error prior to haplotype inference (Holzschuh et al., 2024). Nanopore-specific error profiles were addressed using a denoising pipeline described below.

We inferred per-marker microhaplotypes using a nanopore-adapted version of HaplotypR v.0.5 integrated with DADA2 denoising to correct systematic sequencing errors. DADA2 models error rates from the data and distinguishes true variants, making this combined approach accurate for recovery of multi-SNP haplotypes from short amplicon data despite nanopore error rates (Holzschuh et al., 2024; Holzschuh et al., 2025; Lerch, 2017).

For putative antimalarial resistance-gene orthologues, haplotype consensus sequences were aligned to the PlasmoDB-68 *P. knowlesi* H reference genome and annotated with the corresponding General Feature Format (GFF) to assign nucleotide changes to coding positions and predicted amino-acid substitutions.

Unless otherwise specified, we required ≥10 post-QC reads per microhaplotype. Minor clones were reported only if their within-sample allele frequency (WSAF) was ≥1% of the total read count for that sample and marker (and in all cases supported by ≥10 reads). Sample barcodes were matched to sample IDs, and reads were assigned to markers by primer matching (allowing up to two mismatches). Chimeric read filters were applied. Loci with persistent off-target mapping or primer-dimer signatures were flagged and excluded from summary statistics.

### Population genetic analysis

To evaluate whether the amplicon panel retained structure observed in WGS data, we used 104 published Malaysian WGS genomes compiled by Turkiewicz et al. (2023): 43 Mf-Pk, 30 Mn-Pk, and 31 Pen-Pk genomes (Supplementary Table 5) (Assefa et al., 2015; Diez Benavente et al., 2019; Divis et al., 2018; Hocking et al., 2020; Pinheiro et al., 2015). Reads were mapped to the PlasmoDB-68 *P. knowlesi* H reference genome with the Burrows-Wheeler Aligner (BWA) v0.7.17 (Li & Durbin, 2009), alignments were processed with SAMtools, and variants were jointly called with bcftools mpileup/call using haploid ploidy (Danecek et al., 2021). Genotype calls with DP < 5 were treated as missing. The genome-wide callset was restricted to biallelic SNPs with site quality ≥ 30, call rate ≥ 80%, and alternate-allele count ≥ 2. After invariant sites were removed, 1,181,969 variable SNPs were retained for the genome-wide PCA. Missing genotypes were mean-imputed by SNP, variants were centered and standardized to unit variance, and principal components were obtained by singular-value decomposition.

For PCA, amplicon marker sequences were extracted from the 104 published Malaysian WGS genomes and combined with the sequencing data from the 23 Thai infections. At positions with more than one alternate allele, each alternate allele was represented as a separate variable for PCA; missing calls were mean-imputed per variable, zero-variance variables were removed, variables were centered and standardized to unit variance, and principal components were obtained by singular-value decomposition.

We then constructed a compact, 30-SNP barcode based on the 505 Malaysian WGS SNPs. Candidate SNPs passing call-rate and allele-frequency filters were ranked separately for three pairwise fixation-index (FST) analyses: Mf-Pk versus Mn-Pk, Mf-Pk versus Pen-Pk, and Mn-Pk versus Pen-Pk. SNPs were selected by iteratively screening across the three rankings until 30 population-informative SNPs were retained, spanning 18 amplicons (Supplementary Table 6). The 104 published Malaysian genomes and 23 Thai infections were analysed jointly at these 30 sites using the preprocessing described above. This strategy was conceptually analogous to reduced malaria barcodes built from differentiated loci (Argyropoulos et al., 2023; Wu et al., 2026). Although markers with putative links to drug resistance are commonly excluded from population-genetic analyses, the three drug-resistance-associated loci retained in the panel (*pkdhfr-ts*, *pkcrt*, and *pkmdr1*) showed intermediate heterozygosity (He = 0.67–0.70) comparable to the panel-wide median and were retained in downstream relatedness analyses.

The 28 Peninsular Malaysian and 23 Thai infections were analysed jointly; missing genotypes were mean-imputed by SNP, genotypes were centered without variance standardization, and principal components were obtained by singular-value decomposition. Thai cluster labels were fixed from the discriminant analysis of principal components (DAPC) before this comparison (Jombart et al., 2010).

### Diversity and population genetic analyses

For the Thai sequencing data, multiplicity of infection (MOI) and effective within-host diversity were estimated using MOIRE, which jointly estimates allele frequencies, MOI, and within-host relatedness from polyallelic *Plasmodium* genotypes and reports effective MOI (eMOI) (Murphy & Greenhouse, 2024). eMOI was used as the primary within-host diversity summary. As a sensitivity analysis, we also applied the ranked multi-allelic locus heuristic in dcifer (getCOI, lrank = 2), MOI as the allele count supported by at least two loci. Locus-level expected heterozygosity (He) was calculated from allele frequencies estimated with dcifer::calcAfreq. Two loci that were monomorphic among the Thai infections (PKNH_1019900 and PKNH_1257700) were excluded from He and identity-by-descent (IBD) analyses, leaving 25 polymorphic loci. Pairwise relatedness was inferred with dcifer’s identity-by-descent estimator (ibdDat/ibdPair) (Gerlovina et al., 2022). Population structure was summarized with DAPC (Jombart et al., 2010). BIC was examined across candidate values of K, but K = 2 was retained as a parsimonious first-order summary of the dominant partition rather than selected as the global minimum-BIC solution. The two DAPC groups were defined before comparison with the published Malaysian genomes. Associations between group membership and sampling province or collection year were examined with Fisher’s exact tests, treated as exploratory given small cell counts (Supplementary Table 8).

## Results

### Protocol validation with Thai clinical infections

The final multiplex panel consisted of thirty markers, spanning 13 of the 14 nuclear chromosomes (Supplementary Tables 3 and 4; Supplementary Figure S2). ddPCR quantification of 28 clinical infections from Thailand yielded densities ranging from 7 to 10,530 genome copies/µL of template DNA with a median of 199.5 (Supplementary Table 1). Five infections had parasite densities <10 genome copies/µL. Across all 28 sequenced infections and 30 markers, 4,830,590 reads were obtained from demultiplexed infection libraries (mean 172,521 per infection; excluding 199,466 unclassified reads), of which 2,981,816 met the ≥Q20 filter (mean 106,493; mean Q 25.2; Supplementary Table 9; per-infection read yields are given in Supplementary Table 10; Supplementary Figure S3).

Of the 30 loci, 27 (90%) generated sequence reads in at least one clinical infection. The number of loci successfully recovered per infection increased with parasitemia (Figure 1). The three loci that generated no reads in any infection included PKNH_1429900 (*pkdhps*, a putative drug-resistance marker targeting dihydropteroate synthase) and two microhaplotype loci; all three were excluded from downstream analyses. Five infections yielded data for fewer than eight loci and were excluded from downstream diversity and relatedness analyses. Among the 23 infections retained for downstream diversity and relatedness analyses, a median of 18 of 27 loci were recovered (range: 8-25). For each marker data were obtained for a median of 19 infections (range: 2-23). Across the 27 markers, the per-locus median read depth across infections with reads ranged from 13 to 20,622 reads, and 471 of 756 (62.3%) infection–locus combinations across all 28 infections met the ≥10-read threshold used for haplotype calling (Figure 2, Supplementary Figure S4).

**Figure 1.**
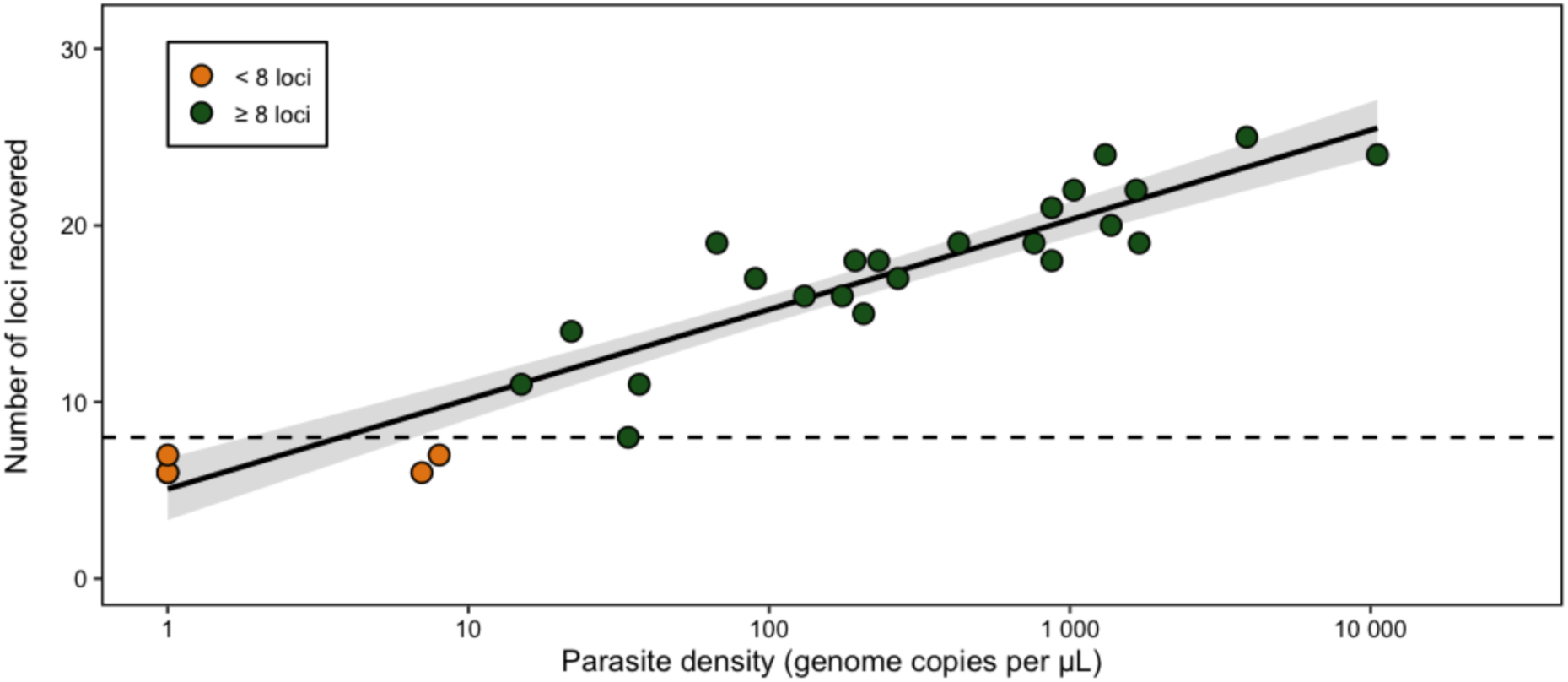
Relationship between parasite density and markers successfully genotyped. The number of loci successfully genotyped per infection increased with parasite density, with a strong linear relationship (R² = 0.88). The dashed line indicates the eight-locus threshold used for downstream analyses; orange points indicate infections below this threshold that were excluded from downstream analyses.

**Figure 2.**
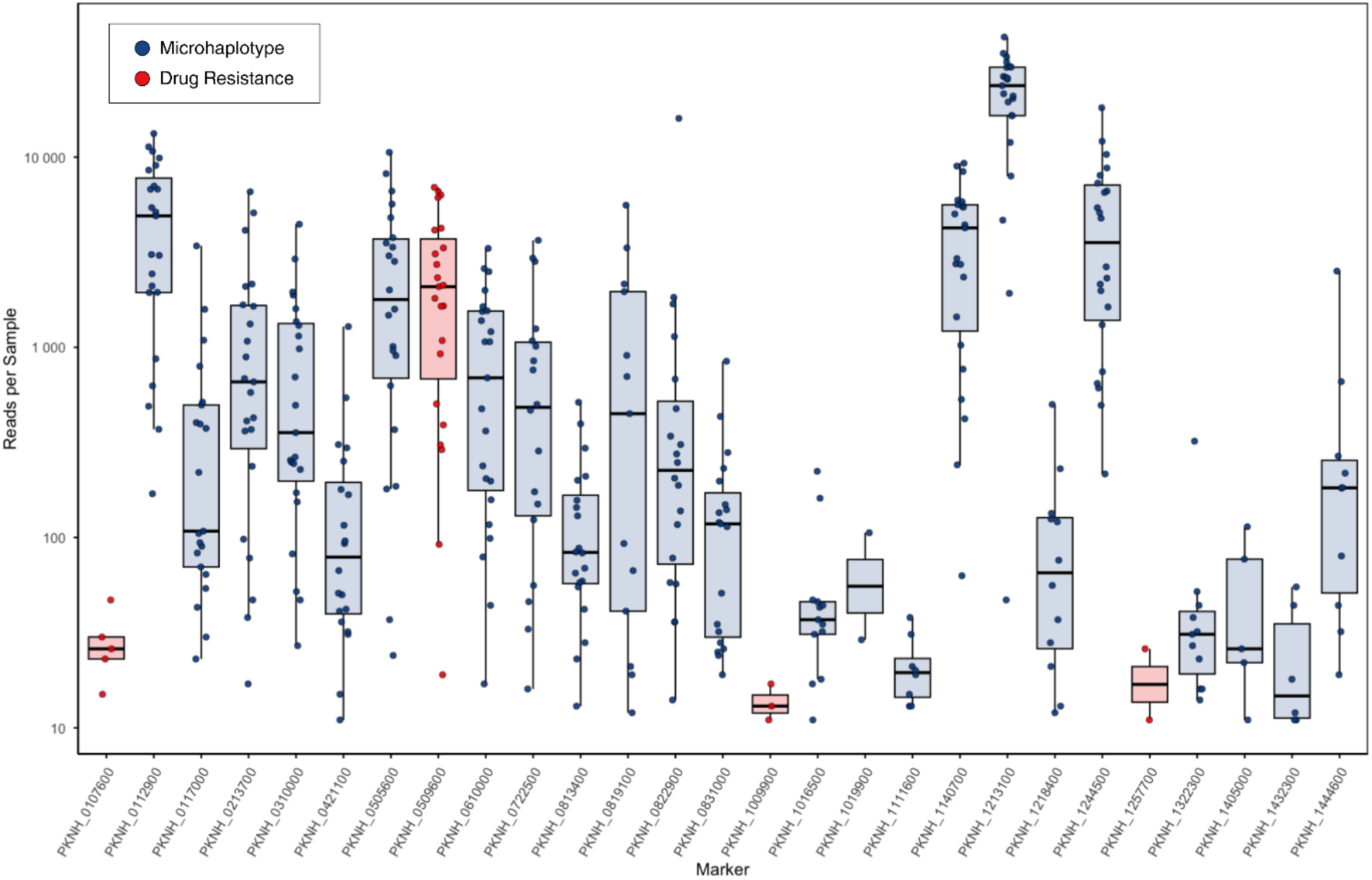
Reads per sample per marker. Boxplots show the distribution of read counts per infection for each marker (log₁₀ scale), with points representing individual infections. Loci targeting putative orthologs of known *P. falciparum* and *P. vivax* drug-resistance genes are shown in red, and microhaplotype loci in blue.

Across the 27 loci, allelic richness ranged from 1 to 8 distinct haplotypes (median = 4; Supplementary Table 11). Two loci were monomorphic: the putative antimalarial resistance-gene orthologue *pkk13* (PKNH_1257700), and a CPSF domain-containing protein (PKNH_1019900). These loci were excluded from downstream diversity and relatedness analyses of the Thai amplicon data but remained included in the WGS analyses.

### Population structure retained across recovered amplicon markers

Genome-wide PCA of the 104 published Malaysian WGS genomes based on 1,181,969 variable SNPs reproduced the published Mf-Pk, Mn-Pk, and Pen-Pk separation (Figure 3A). Within the 27 recovered amplicon markers, the Malaysian WGS genomes contained 505 variable SNPs. The Thai infections contained 204 variant positions, of which 123 were shared with the Malaysian dataset, yielding a combined 586 SNPs (382 Malaysia-only and 81 Thai-only; Supplementary Figure S5). PCA was recalculated jointly across the 104 Malaysian WGS genomes and 23 Thai infections at these 586 positions. The joint PCA retained the broad Malaysian population structure and placed the Thai infections within or adjacent to Pen-Pk space (Figure 3B).

**Figure 3.**
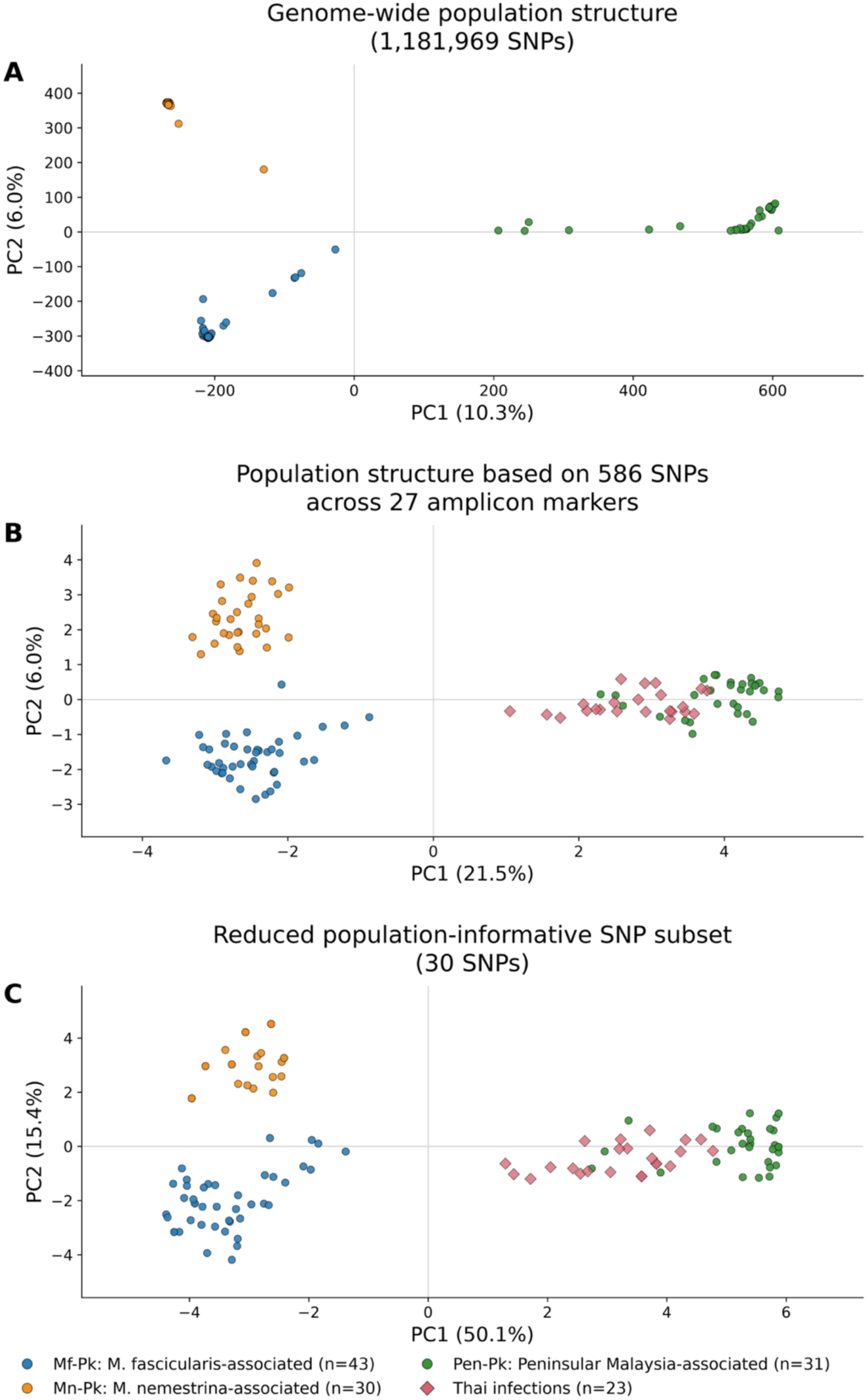
Comparison of regional *P. knowlesi* population structure using whole-genome and targeted single-nucleotide polymorphism (SNP) datasets. (A) Whole-genome sequencing (WGS)-based principal component analysis (PCA) of 104 published Malaysian WGS genomes using 1,181,969 variable SNPs. (B) Joint PCA of the 104 published Malaysian WGS genomes and 23 Thai infections using 586 validated positions across the 27 ddPCR- AmpSeq amplicon markers. (C) Joint PCA of the same genomes and infections using a 30-SNP population-informative subset selected from the 505 Malaysian WGS SNPs to summarize differentiation among *M. fascicularis*- associated *P. knowlesi* (Mf-Pk), *M. nemestrina*-associated *P. knowlesi* (Mn-Pk), and Peninsular Malaysia-associated *P. knowlesi* (Pen-Pk). Thai infections were analyzed jointly with the published Malaysian WGS genomes in panels B and C. Population labels, accessions, and source studies follow Turkiewicz et al. (2023; Supplementary Table 5).

The reduced 30-SNP barcode provided a compact representation of the same broad differentiation while retaining the Thai-Pen-Pk affinity (Figure 3C; Supplementary Table 6). The larger fraction of variance captured by PC1 in this selected-SNP analysis reflects deliberate enrichment for differentiated sites and is not interpreted as superiority to the genome-wide analysis.

Fine-scale Pen-Pk structure was evaluated using the 28 published Peninsular Malaysian clinical infections analyzed by Hocking et al. (2020), for which WGS defined three subclusters: Pen A (n = 15), Pen B (n = 10), and Pen C (n = 3) (Supplementary Table 2). Among the 586 positions identified across the Malaysian genomes and Thai AmpSeq data, 239 positions were monomorphic among these 28 infections. Due to insufficient sequencing depth in the Malaysian WGS data or the Thai AmpSeq data, 17 markers containing 234 diverse SNPs were retained for the PCA analysis. In the fine-scale Pen-Pk comparison, Pen C remained separated from the Pen A/B region, whereas Pen A and Pen B overlapped substantially (Figure 4A).

**Figure 4.**
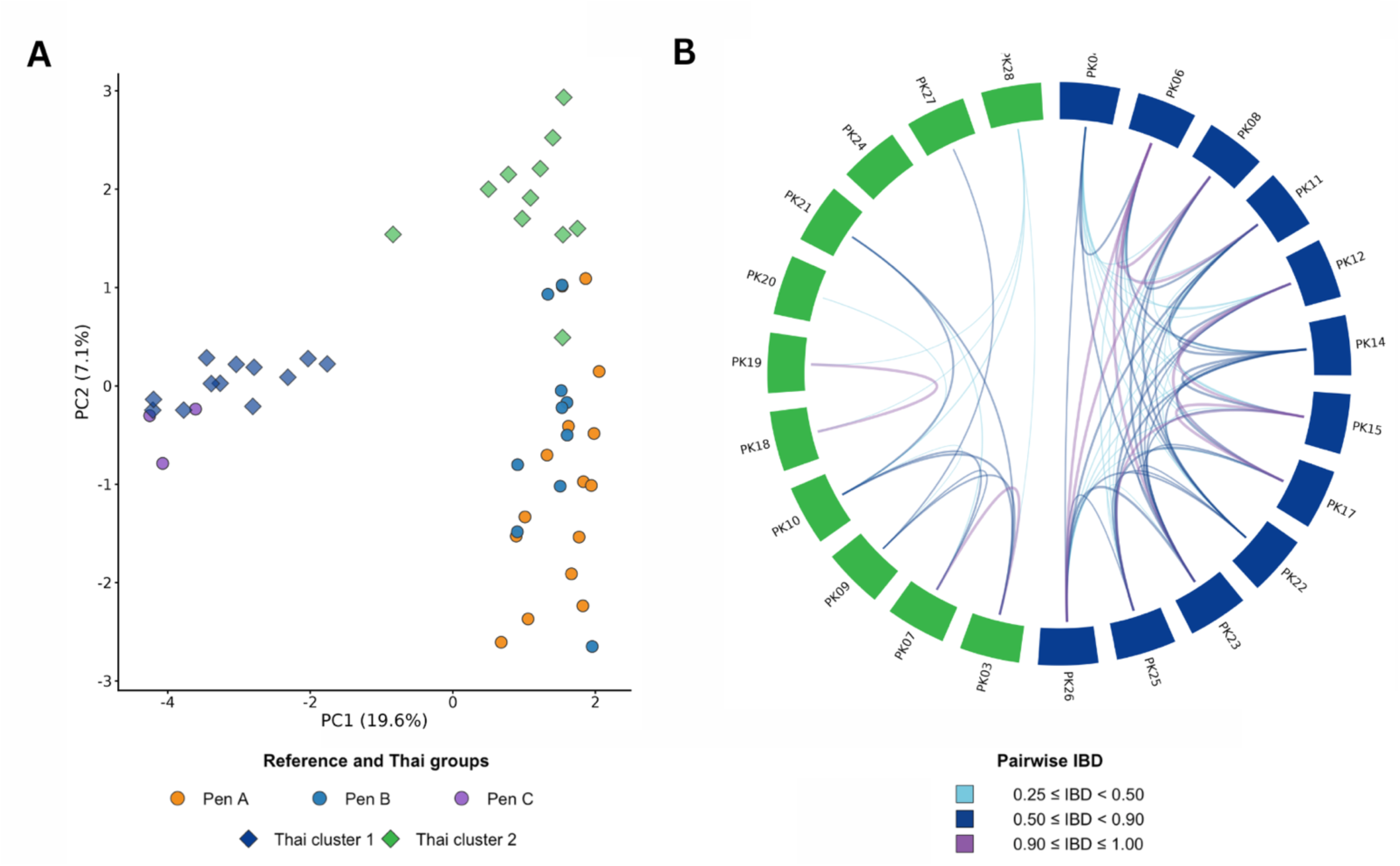
Thai *P. knowlesi* relatedness and fine-scale comparison with published Peninsular Malaysian clinical infections. (A) Joint PCA based on 234 SNPs retained for the combined analysis of 28 published Peninsular Malaysian clinical infections and 23 Thai infections. Published Peninsular Malaysian infections are colored by the Pen A, Pen B, and Pen C designations, and Thai infections are shown as diamonds colored by Thai DAPC group. PC1 and PC2 explain 19.6% and 7.1% of the variance, respectively. (B) Pairwise identity-by-descent (IBD) among the 23 Thai infections, arranged by the two groups defined independently by discriminant analysis of principal components (DAPC). Links indicate 0.25 ≤ IBD < 0.50, 0.50 ≤ IBD < 0.90, and 0.90 ≤ IBD ≤ 1.00.

### Thai *P. knowlesi* diversity and relatedness

eMOI classified all 23 Thai infections as effectively monoclonal (eMOI = 1.0; Supplementary Table 12). Across the 25 polymorphic loci, within-marker genetic diversity was moderate (mean He = 0.69, median He = 0.70, range 0.22–1.00). Four markers showed high diversity (He ≥ 0.80). The three loci with putative links to drug resistance showed intermediate diversity: the bifunctional dihydrofolate reductase-thymidylate synthase gene (*pkdhfr-ts*; PKNH_0509600, He = 0.69), the chloroquine resistance transporter (*pkcrt*; PKNH_0107600, He = 0.70), and multidrug resistance protein 1 (*pkmdr1*; PKNH_1009900, He = 0.67). Per-locus He values are provided in Supplementary Table 13.

DAPC identified a clear split in the Thai dataset, defining Thai cluster 1 (n = 12) and Thai cluster 2 (n = 11). Both Thai clusters included infections from multiple provinces and collection years. Cluster membership was not significantly associated with province (Fisher’s exact test, p = 0.2105) or collection year (p = 0.2423; Supplementary Table 8). When compared with the previously described Pen A/B/C clusters in Peninsular Malaysia, Thai cluster 2 was positioned within or near the Pen A/B region, whereas Thai cluster 1 was positioned toward Pen C (Figure 4A).

We then used pairwise IBD to examine recent shared ancestry. Across 253 pairwise comparisons among the 23 Thai infections, mean IBD was 0.195. Forty-six pairs (18.2%) had IBD ≥0.50 and 11 pairs (4.3%) had IBD ≥0.90, including 10 pairs with IBD = 1.0 (Figure 5C; Supplementary Table 14). All 77 pairs with IBD ≥0.25 occurred within the two DAPC groups (Figure 4B and Figure 5A). At IBD ≥0.50, the threshold network comprised two main connected components plus a two-infection pair (Figure 5B). Per-locus haplotype networks also showed group-enriched haplotypes at multiple loci (Supplementary Figure S6).

**Figure 5.**
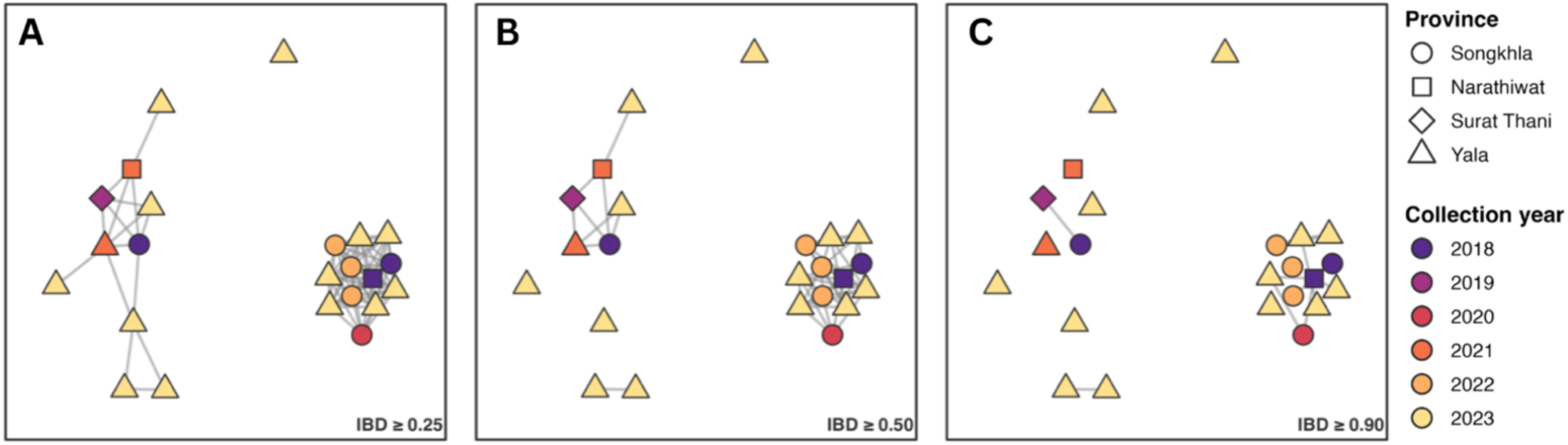
Identity-by-descent (IBD) networks among Thai *P. knowlesi* infections by sampling province and collection year. Pairwise IBD networks among the 23 retained Thai infections are shown at thresholds of (A) IBD ≥ 0.25, (B) IBD ≥ 0.50, and (C) IBD ≥ 0.90. Nodes represent individual infections, with shape indicating sampling province and color indicating collection year. Edges connect infection pairs with IBD values meeting or exceeding the indicated threshold.

### Putative antimalarial resistance-gene orthologue variants among Thai infections

Four loci were examined for coding variation: *pkcrt* (PKNH_0107600), *pkdhfr-ts* (PKNH_0509600), *pkmdr1* (PKNH_1009900), and *pkk13* (PKNH_1257700). Nucleotide changes, codon positions, amino-acid substitutions, infection-level haplotype support, and literature context are provided in Supplementary Table 15. No coding variant was detected in *pkcrt*. Nonsynonymous changes were observed in *pkdhfr-ts* and *pkmdr1*: L4F and E6K were each detected in one infection, G32V in five infections, and R34L in 14 infections across four recovered haplotypes; *pkmdr1* N1011S was detected in one infection. The synonymous *pkk13* G531G substitution was detected in two infections, and no nonsynonymous *pkk13* change was detected in the amplicon.

## Discussion

A panel of 27 amplicon markers retained the major population structure seen in the published WGS data. The panel separated infections from Borneo and Peninsular Malaysia, and among Bornean infections distinguished infections associated with long-tailed and pig-tailed macaques. The 30-SNP subset provided a compact representation of the same broad Mf-Pk/Mn- Pk/Pen-Pk differentiation. Because the full amplicon sequences retain substantially more variation, the panel also supports focused epidemiological investigations, where distinguishing individual infections is important for investigating local transmission.

Among the 23 Thai infections, DAPC and IBD supported two genetic groups with different positions in the published Peninsular Malaysian PCA space (Figure 4; Hocking et al., 2020). At the broader scale, both groups showed affinity to Pen-Pk rather than the Bornean Mf-Pk or Mn-Pk populations (Divis et al., 2015; Divis et al., 2018; Turkiewicz et al., 2023). These groups might reflect the structure observed in peninsular Malaysia, where three groups were observed. Of note, this structure was based on only 28 genomes, and one group included only three infections (Hocking et al., 2020).

Both Thai genetic groups contained infections from multiple provinces and each spanned multiple collection years. Broader sampling of humans, macaques, and vectors across Thailand and Peninsular Malaysia will be necessary to evaluate parasite movement between countries or establish transmission direction. Thai infections carried nonsynonymous variation in *pkdhfr-ts* and *pkmdr1*, including frequent *pkdhfr-ts* R34L. Variation in orthologues of markers of drug resistance might serve as an indicator for exposure to drug in human hosts followed by onward transmission (Grigg et al., 2016; Putaporntip et al., 2026; Saif, 2023; Sangsri et al., 2023), yet phenotypic or functional evidence is still required before individual substitutions are interpreted as resistance determinants.

In conclusion, this study establishes a *P. knowlesi* targeted-sequencing framework that links three levels of information: first-order structure and recent relatedness within field infections, broad affinity to WGS-defined Mf-Pk/Mn-Pk/Pen-Pk populations, and finer affinity within Pen-Pk. The coexistence of two genetically differentiated Pen-Pk-like groups across provinces and collection years suggests that parasite structure in southern Thailand is not explained by simple geographic distances and may reflect broader ecological or cross-border processes that will require coordinated sampling of human, macaque, and vector populations to resolve.

## Acknowledgements

This research was supported by Mahidol University through the National Science Research and Innovation Fund (NSRF), the National Institute of Allergy and Infectious Diseases (NIAID), National Institutes of Health (NIH), USA, under award U19AI181583, and the University of Notre Dame Asia Research Collaboration Grant (2023–2024) to C. Koepfli in partnership with Mahidol University, Bangkok, Thailand.

## Ethics Approval and Patient Consent

This study was approved by the Ethics Committee of the Faculty of Tropical Medicine, Mahidol University, Bangkok (TMEC 18-009 and TMEC 24-109) and the University of Notre Dame Institutional Review Board (23-09-8102). All participants provided informed consent to participate. All laboratory analyses used de-identified DNA archived at the Mahidol Vivax Research Unit, Bangkok, Thailand.

## Data Accessibility and Benefit-Sharing

## Data Accessibility Statement

Raw nanopore sequence data generated in this study are deposited in the European Nucleotide Archive under project accession PRJEB115548; per-sample accessions are ERS30643652– ERS30643679. The 28 published Peninsular Malaysian clinical infections used for target discovery and fine-scale comparison and the 104 published Malaysian WGS genomes used for regional comparison derive from published whole-genome data described by Hocking et al. (2020), Turkiewicz et al. (2023), and the underlying studies, with genome identifiers listed in Supplementary Tables 2 and 5. Thai infection metadata and the 30-SNP population-informative subset are provided in Supplementary Tables 1 and 6, respectively. The complete supplementary tables, processed genotype datasets, and analysis scripts are publicly available through Zenodo at https://doi.org/10.5281/zenodo.23406811.

## Benefit-Sharing Statement

Benefits generated by this research include a novel assay for genomic surveillance that can be used by Thai and other scientists and malaria control programs. The sequencing assay was taught to scientists from Thailand and neighbouring countries in a workshop at Mahidol University jointly run by scientists from the University of Notre Dame and Mahidol University. Thai scientists are included as co-authors on this publication.

## Author Contributions

Conceptualization: GS, PT, JS, WN, CK. Methodology: PT, JS, WN, CK. Investigation: GS, PT. Formal analysis: GS, AH. Resources: PT, JS, WN, CK. Writing – original draft: GS, CK. Writing – review & editing: GS, PT, AH, JS, WN, CK. Supervision: CK.

---

**Supplementary Figure S1.**
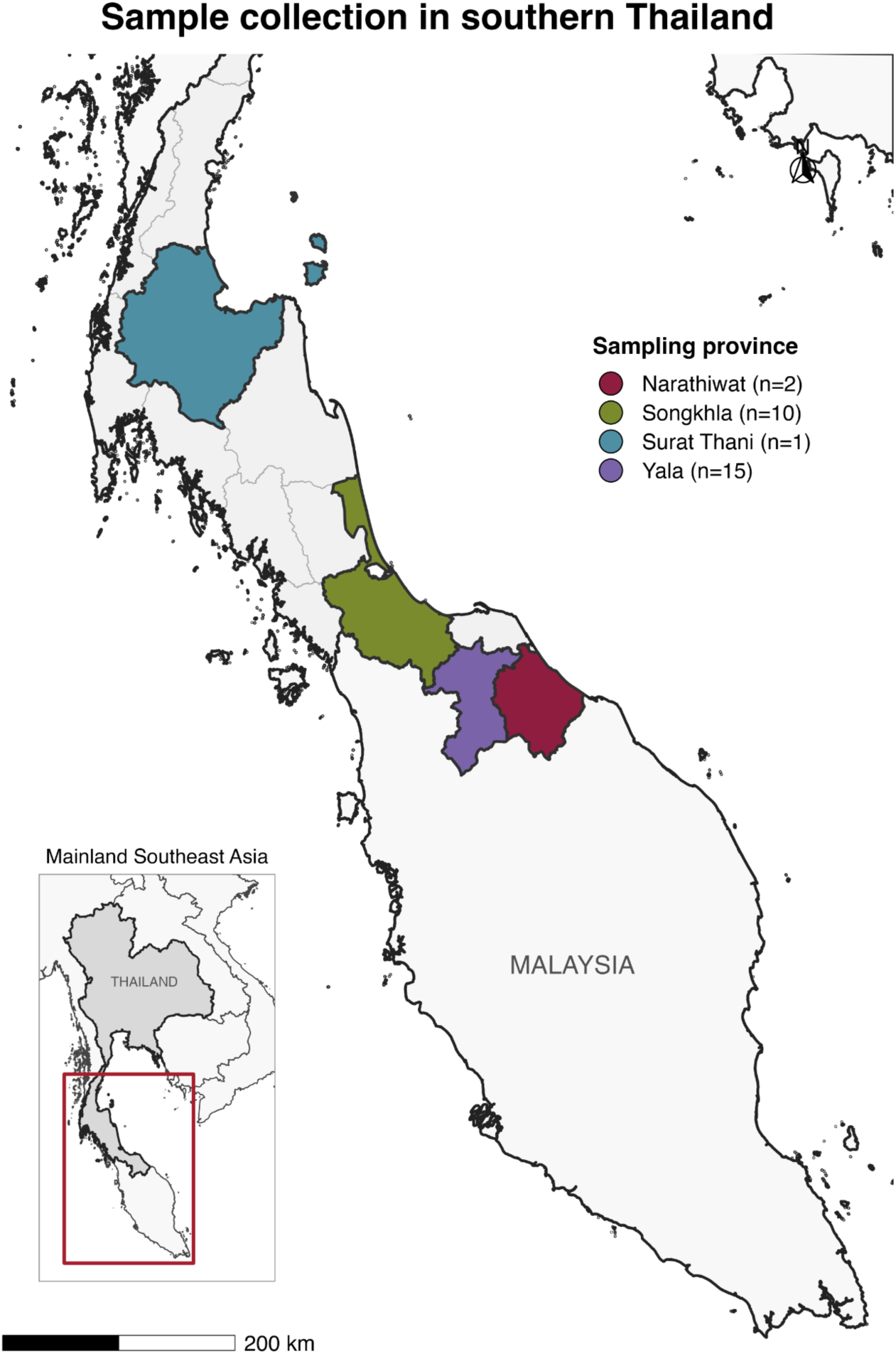
Clinical infections sampled in southern Thailand. Map showing the four provinces represented among all 28 sequenced clinical infections: Narathiwat (n = 2), Songkhla (n = 10), Surat Thani (n = 1), and Yala (n = 15). The inset places the study area within mainland Southeast Asia. Province counts refer to the full sequenced infection set; the 23 infections retained for population- genetic analyses are detailed in Supplementary Table 1.

**Supplementary Figure S2.**
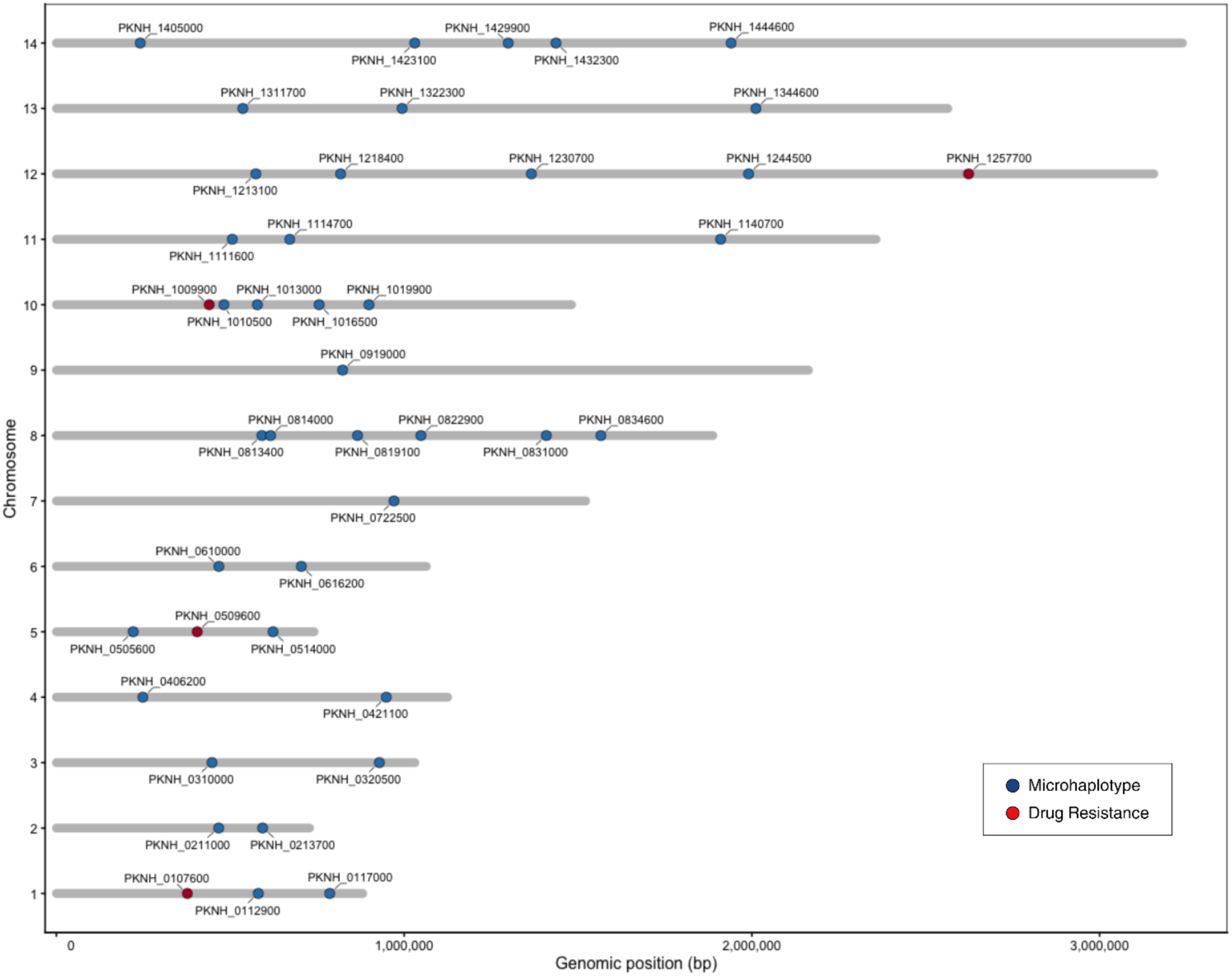
Chromosomal distribution and genomic coordinates of the 43 initial candidate amplicons screened during panel development. The candidate set included microhaplotype regions selected from the Peninsular Malaysian WGS diversity screen together with putative antimalarial resistance-gene orthologues and spanned all 14 *P. knowlesi* chromosomes. Thirty primer pairs passed singleplex screening and formed the final experimental multiplex panel; because the only chromosome 9 candidate (PKNH_0919000) failed screening, the final 30-locus panel represents 13 of 14 chromosomes. The Peninsular Malaysian WGS diversity screen used for microhaplotype target discovery was derived from Hocking et al. (2020) (Supplementary Table 2).

**Supplementary Figure S3.**
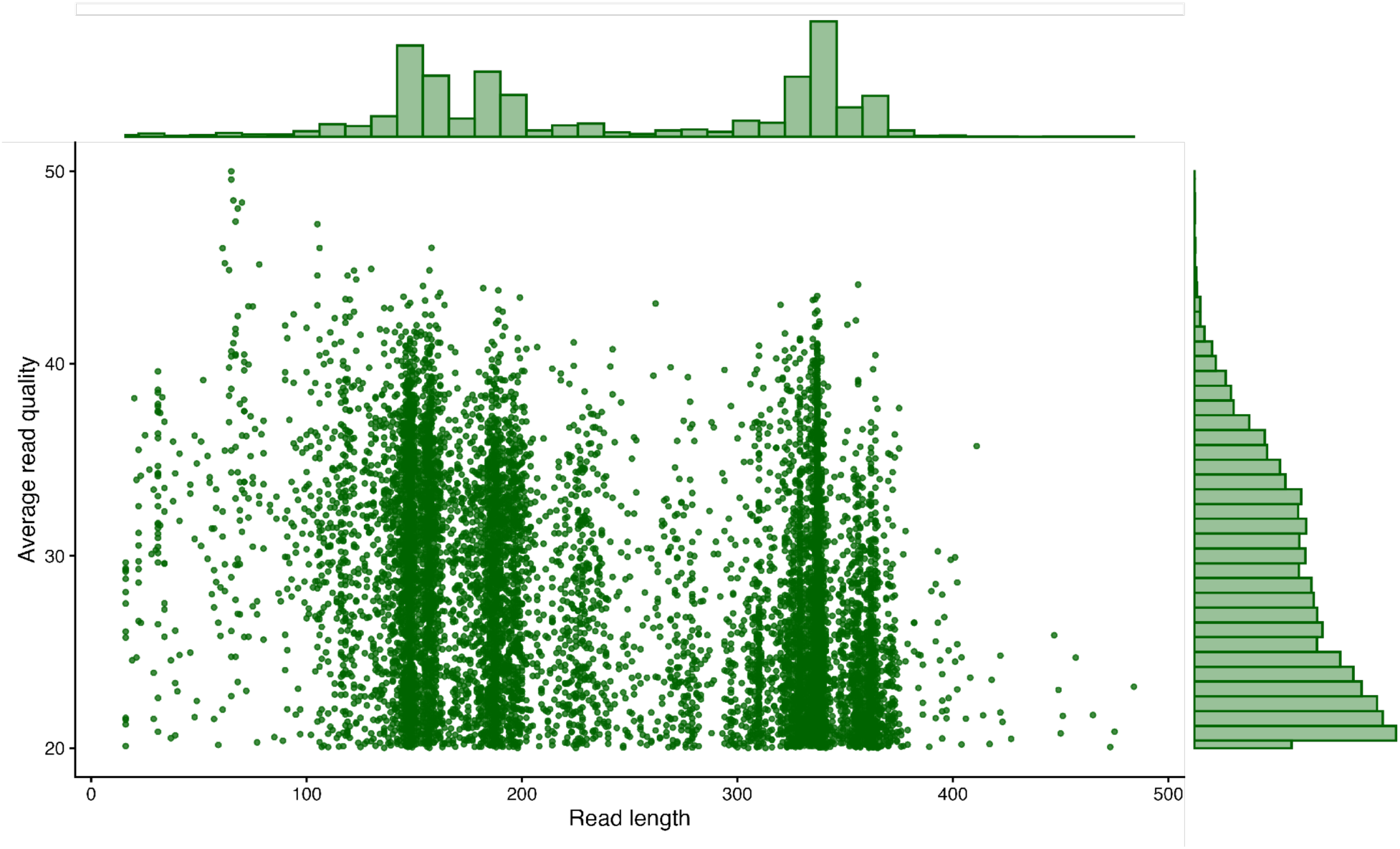
Read length and basecall quality distribution for the nanopore AmpSeq run. Scatter plot showing read length versus mean basecall quality, with marginal histograms summarizing the distribution of read lengths and quality scores. The plot was reconstructed from the embedded Plotly data in the NanoPlot HTML report.

**Supplementary Figure S4.**
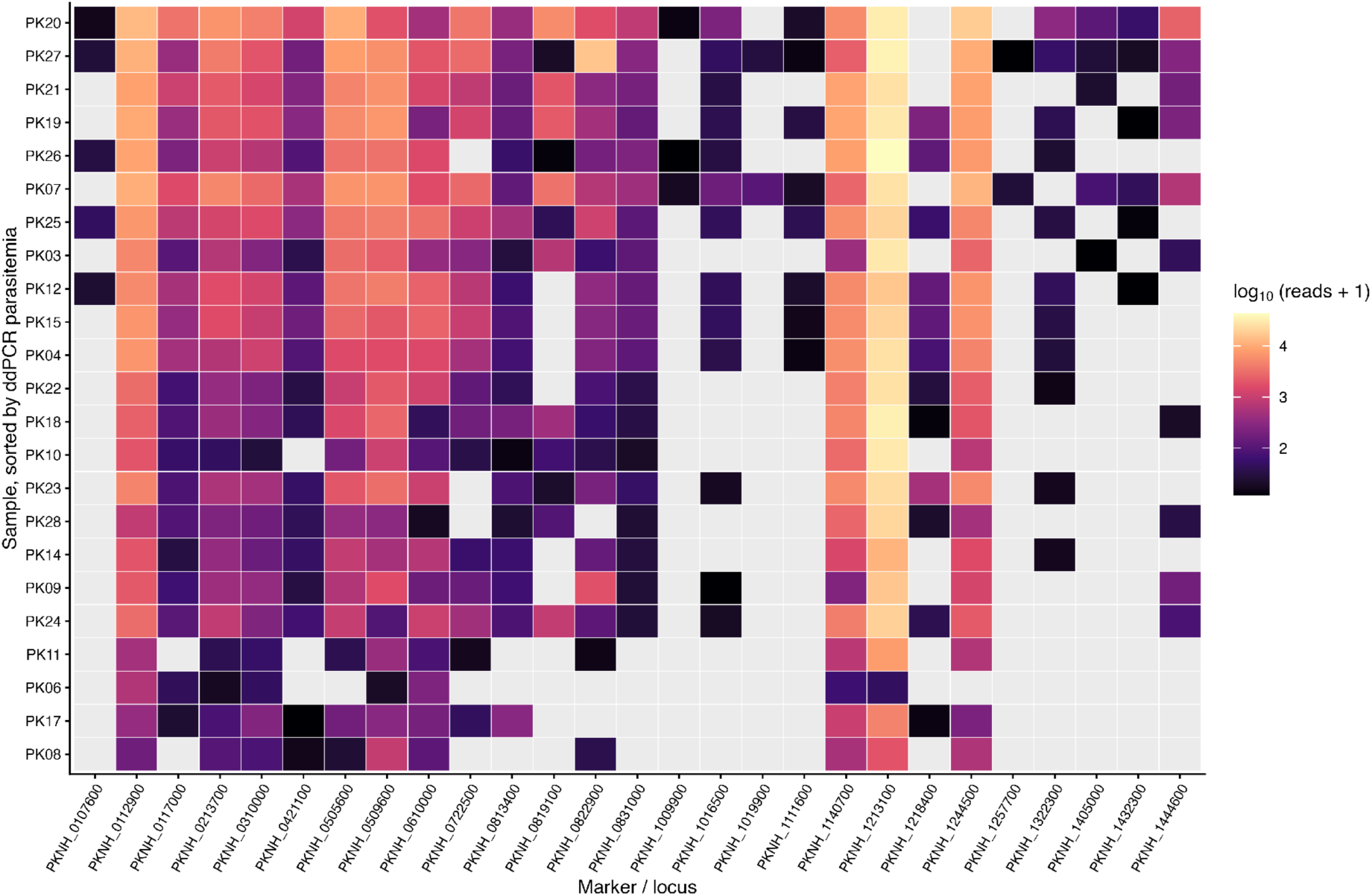
Infection-by-locus read recovery across sequenced *P. knowlesi* infections. Heatmap showing summed haplotype-supporting reads per infection and marker from the HaplotypR output. Read depth was calculated by summing reads across retained haplotypes for each infection- marker combination. Grey cells indicate markers with no retained haplotype call.

**Supplementary Figure S5.**
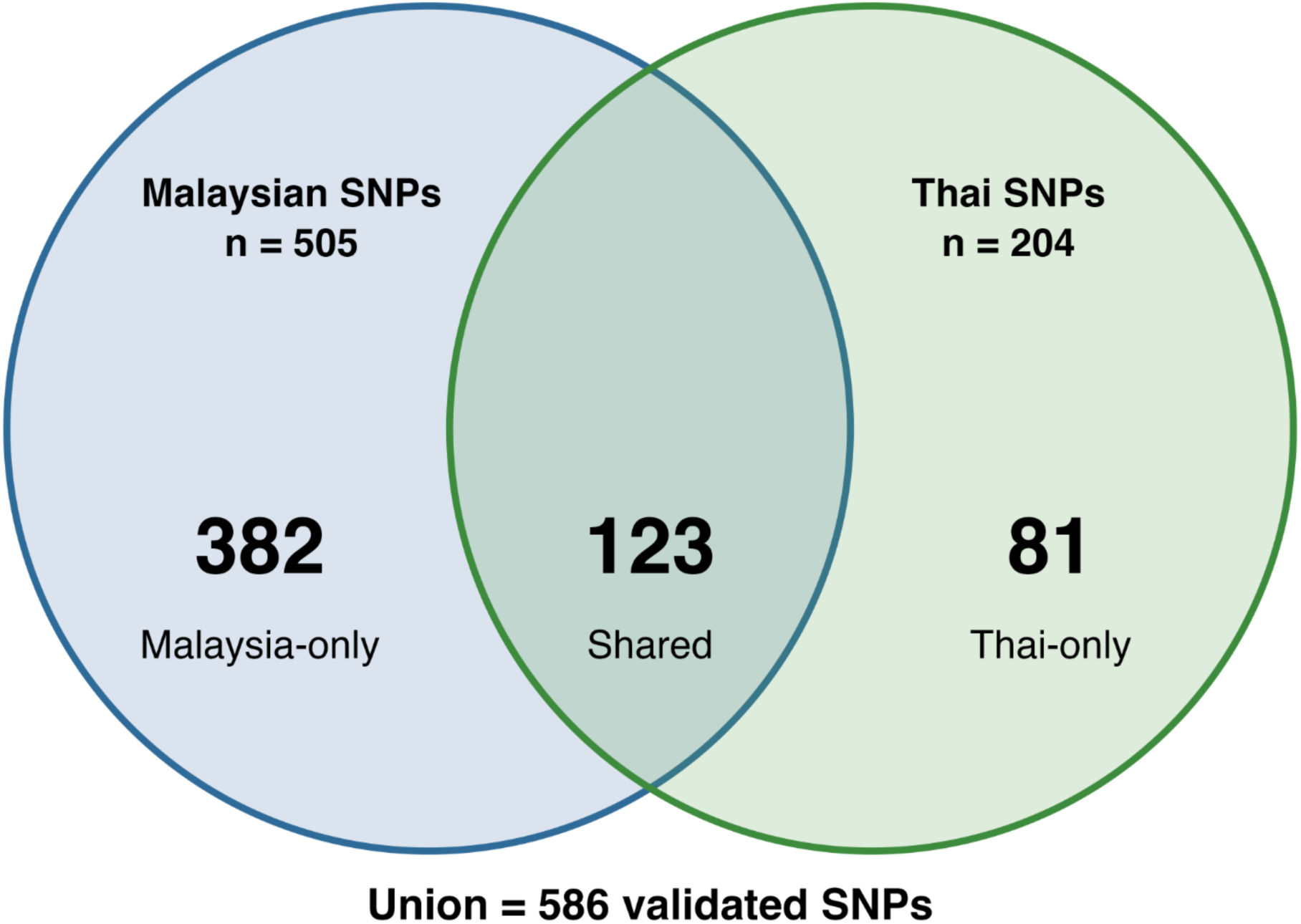
Overlap between Malaysian and Thai variant positions across the 27 recovered amplicon markers. The published Malaysian WGS dataset contained 505 variable SNPs within the recovered amplicon markers, while 204 variant positions were identified among the Thai infections relative to the *P. knowlesi* H reference genome. Of these, 123 positions were shared, 382 occurred only among the published Malaysian WGS genomes, and 81 occurred only among Thai infections, yielding a combined 586 SNPs. The 204 Thai variant positions comprised 176 positions segregating among Thai infections and 28 positions fixed for a non-reference allele.

**Supplementary Figure S6.**
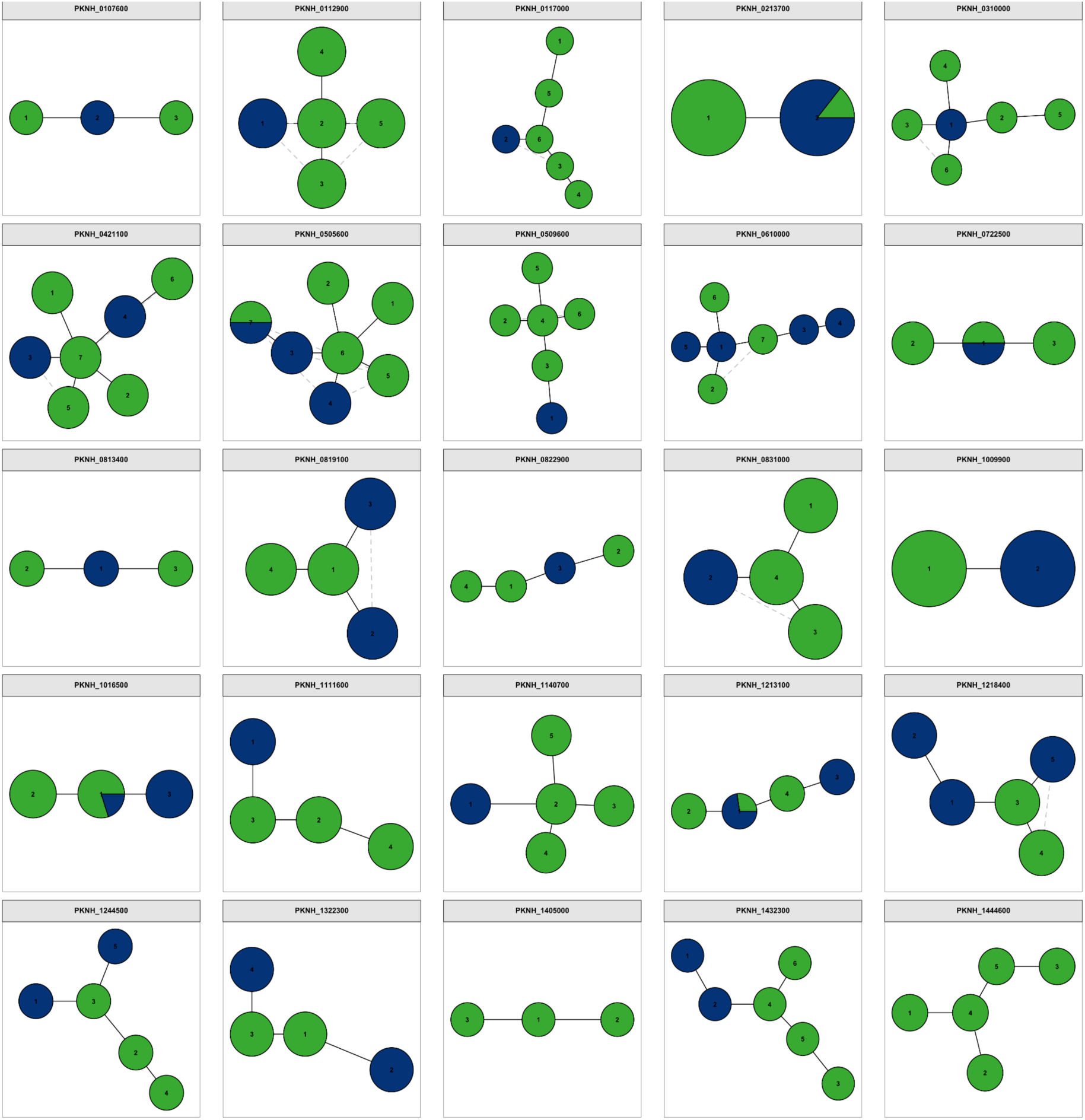
Haplotype networks for the 25 polymorphic loci included in downstream analyses. Each panel represents one locus, and each node represents one haplotype observed across the 23 analyzed infections. Edges indicate mutational steps. Numbers within nodes correspond to haplotype identifiers as inferred by the nanopore-adapted HaplotypR pipeline.

